# Y380Q novel mutation in receptor-binding domain of SARS-CoV-2 spike protein together with C379W interfere in the neutralizing antibodies interaction

**DOI:** 10.1101/2021.09.10.21262695

**Authors:** Ivaine Tais Sauthier Sartor, Fernanda Hammes Varela, Mariana Rost Meireles, Luciane Beatriz Kern, Thaís Raupp Azevedo, Gabriela Luchiari Tumioto Giannini, Mariana Soares da Silva, Meriane Demoliner, Juliana Schons Gularte, Paula Rodrigues de Almeida, Juliane Deise Fleck, Gabriela Oliveira Zavaglia, Ingrid Rodrigues Fernandes, Caroline Nespolo de David, Amanda Paz Santos, Walquiria Aparecida Ferreira de Almeida, Victor Bertollo Gomes Porto, Marcelo Comerlato Scotta, Gustavo Fioravanti Vieira, Fernando R. Spilki, Renato T. Stein, Márcia Polese-Bonatto, COVIDa study group

## Abstract

**Background:** The emergence of SARS-CoV-2 variants is a current public health concern possibly impacting COVID-19 disease diagnosis, transmission patterns and vaccine effectiveness.

**Objectives:** To describe the SARS-CoV-2 lineages circulating early pandemic among samples with *S* gene dropout and characterize a novel mutation in receptor-binding domain (RBD) of viral spike protein.

**Study design:** Adults and children older than 2 months with signs and symptoms of COVID-19 were prospectively enrolled from May to October 2020 in Porto Alegre, Brazil. All participants performed RT-PCR assays for diagnosing SARS-CoV-2, samples with *S* gene dropout and Ct < 30 (cycle threshold) were submitted to whole genome sequencing (WGS), and homology modeling and physicochemical properties analysis were performed.

**Results:** 484/1,557 participants tested positive for SARS-CoV-2. The *S* gene dropout was detected in 7.4% (36/484) as early as May, and a peak was observed in early August. WGS was performed in 8 samples. The B.1.1.28, B.1.91 and B.1.1.33 lineages were circulating in early pandemic. The RBD novel mutation (Y380Q) was found in one sample occurring simultaneously with C379W and V395A, and the B.1.91 lineage in the spike protein.

**Conclusion:** Mutations in the SARS-CoV-2 spike region were detected early in the COVID-19 pandemic in Southern Brazil, regarding the B.1.1.28, B.1.91 and B.1.1.33 lineages identified. The novel mutation (Y380Q) with C379W, modifies important RBD properties, which may interfere with the binding of neutralizing antibodies (CR3022, EY6A, H014, S304).

**Highlights:**

- Characterization of novel mutation (Y380Q) in RBD of SARS-CoV-2 spike protein
- The Y380Q and C379W modify important properties in the SARS-CoV-2 RBD region
- The RBD mutations may interfere with the binding of neutralizing antibodies
- The B.1.1.28, B.1.91 and B.1.1.33 lineages were circulating in early pandemic

## Introduction

SARS-CoV-2 is a single RNA-stranded virus with high mutation rates. Strategies to mitigate the pandemic include the knowledge of its viral genome and expected mutations. These features could impact disease severity, virus transmission, and vaccine strategies [1–3]. As the COVID-19 pandemic evolves, there has been concern about the emergence of new SARS-CoV-2 mutations in the receptor binding domain (RBD) from the *S* region, due to probable effects on both virus transmissibility and the generation of escape mutants from antibodies previously formed to heterologous lineages and vaccines [4].

Genetic alterations in the RBD of SARS-CoV-2 may improve the affinity of the virus to binding host cells and these changes may lead to higher transmission rates [5,6]. Binding affinity of the S protein and ACE2 make this region a key target for potential therapies and diagnosis [7]. COVID-19 molecular diagnostic tests directed to the *S* gene use it as one of the RT-PCR multiple target-regions.

Our aim was to measure the prevalence of the *S* dropout and characterize the SARS-CoV-2 mutations in the RBD region in a cohort during the early pandemic.

## Materials and Methods

### 1. Participants’ selection

A prospective cohort study enrolled adults and children seeking care at emergency rooms, outpatient clinics, or hospitalized in general wards or intensive care units (ICU) at Hospital Moinhos de Vento and Hospital Restinga e Extremo Sul, both in Porto Alegre, Brazil. From May to early October 2020 were included participants presenting signs or symptoms suggestive of COVID-19 (cough, fever, or sore throat). The key exclusion criteria was a negative SARS-CoV-2 RT-PCR result or failure to sample collection. The study was performed in accordance with the Decree 466/12 of the National Health Council [8] and Clinical Practice Guidelines, after approval by the Hospital Moinhos de Vento IRB nº 4.637.933. All participants included in this study provided written informed consent.

### 2. SARS-CoV-2 detection and sequencing

All participants performed qualitative RT-PCR assay to SARS-CoV-2 detection as described elsewhere [9]. Additionally, *S* gene dropout samples with cycle threshold less than 30 (Ct < 30.0) were submitted to whole genome sequencing (WGS) using the Illumina MiSeq high-throughput. RNA was extracted from oropharyngeal swab samples and the RT reaction was performed using SuperScript IV reverse transcriptase (Thermo Fisher Scientific). Library preparation was conducted using QIAseq SARS-CoV-2 Primer Panel paired for library enrichment and QIAseq FX DNA Library UDI Kit, according to the manufacturer instructions (Qiagen, Hilden, Germany). MiSeq Reagent Kit v3 was used for sequencing (600-cycle). FASTQ reads were imported to Geneious Prime, trimmed (BBDuk 37.25), and mapped against the reference sequence hCoV-19/Wuhan/WIV04/2019 (EPI_ISL_402124) available in EpiCoV database from GISAID [10]. Complete genome alignment was performed with the sequences generated. Fifty-nine Brazilian SARS-CoV-2 complete genomes and the reference sequence (EPI_ISL_402124) (>29 kb) were retrieved from the GISAID database using Clustal Omega. Maximum Likelihood phylogenetic analysis was applied under the General Time Reversible model allowing for a proportion of invariable sites and substitution rates in Mega X applying 200 replicates and 1000 bootstrap.

### 3. Co-localization of Y380Q with B and T-cell epitopes

#### B-cell epitopes

Wild type (Y380) and mutated spike protein sequences (Q380) were submitted to Bepipred 1.0 and 2.0 to detect putative humoral epitopes through HMMs and Random forest algorithms [11,12]. To increase sensitivity we set a threshold of −0.2 (Bepipred 1.0) and 0.45 (Bepipred 2.0).

#### T-cell epitopes

The search in Immune Epitope Database (IEDB) considered T-cells epitopes for SARS-CoV-2 spike protein (region of 10 residues flanking the Y380Q) with 70% similarity in BLAST. Potential binder sequences of representative supertypes MHC-I alleles (HLA-A^*^01:01, HLA-A^*^02:01, HLA-A^*^03:01, HLA-A^*^24:02, HLA-A^*^26:01, HLA-B^*^07:02, HLA-B^*^08:01, HLA-B^*^27:05, HLA-B^*^39:01, HLA-B^*^40-01, HLA-B^*^58:01, HLA-B^*^15:01) were predicted by NetMHCPan-4.1 [13,14].

### 4. Structural modifications and their impact on B and T-cells epitopes recognition

#### Homology modeling

For the wild type template selection the SARS-CoV-2 surface glycoprotein sequence (NCBI accession number: YP_009724390) was submitted to BLAST and SwissModel tools. Template crystal candidates were evaluated by the GMQE, QMEAN, Z-score, and residues distribution in the Ramachandran plot, using ERRAT, PROCHECK, PDBsum, ModFold, SwissModel [15–18]. Protein Data Bank (PDB) 7CWL (3.8Å) was chosen for the approach. Phyre-2 software was employed to homology modeling using the expert mode (one-to-one threading job) for constructing models based on wild protein (P0DTC2) and mutated sequences [19].

#### Physicochemical properties

Electrostatic potential (EP) was verified through Delphi web server calculations and PIPSA [20,21]. The residue exposure characteristics such as hydrophobicity and the Accessible Solvent Surface Area (aSAS) were estimated using the Chimera interface [22].

#### Antibodies interaction

Crystal complexes of the RBD region with antibodies were recovered from PDB and examined to obtain experimental information of structural binding regions. The LigPlot program was applied to infer protein-antibody interaction sites [23]. Hydrogen-bonds (H-bonds) inferences among the RBD domain and antibodies were calculated in the Chimera interface.

### 5. Statistical analysis

Data normality assumptions were verified for continuous variables, and median values and interquartile ranges (IQR) were calculated. Pearson’s Chi-square test was used to evaluate proportions between the identified and undetermined results from *S* gene target, on the epidemiological week; Fisher’s exact test was used to compare the frequencies of *S* dropout considering outpatient and inpatient populations. All analyses were performed in R 3.5.0 statistical software [24].

## Results

A total of 1,557 participants were screened and 484 were detected positive for SARS-CoV-2 (Supplementary Figure 1). Of these, 98 (20.2%) subjects were hospitalized and 386 (79.8%) were seen as outpatients only.

*S* dropout was characterized as undetermined RT-PCR values for the *S* gene target, and detected values for *ORF1ab* and *N* target probes. We observed a total *S* dropout of 36/484 (7.4%), while *ORF1ab* and *N* gene targets showed no dropout (Figure 1). Additionally, an increase in frequency of undetermined results of the *S* gene target was identified on 10^th^ August week with 8/26 dropouts (30.8%, P = 0.007). No differences were observed for other epidemiological weeks. *S* gene dropout was detected in 12/98 hospitalized subjects (12.2%), whereas for outpatients the frequency was 24/386 (6.2%) with an OR (95% CI) of 2.10 (0.92-4.57, P = 0.052).

**Figure 1.**
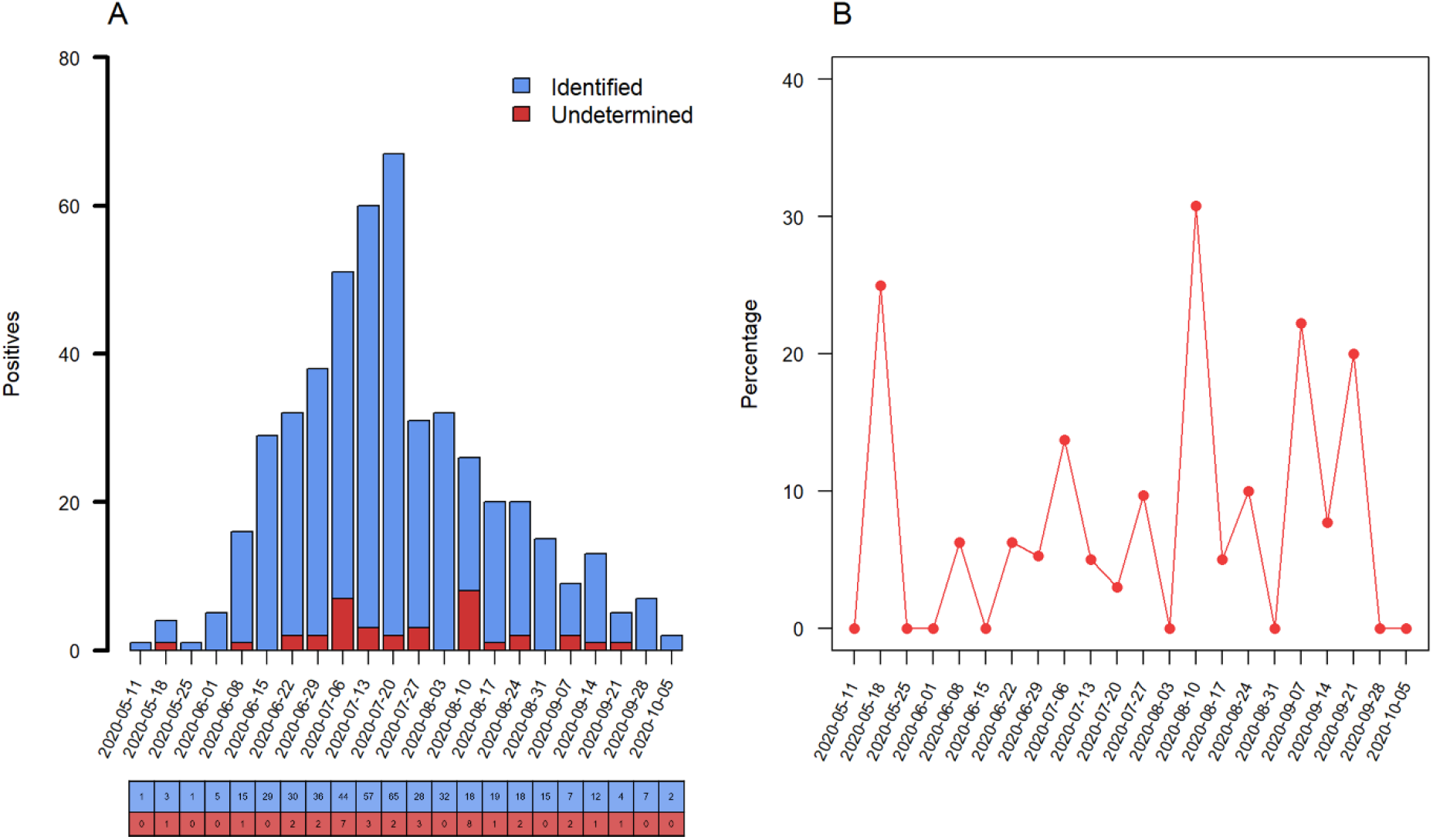
Prevalence of *S* dropout profile from May to October 2020, the dates correspond to the Monday of each epidemiological week. (**A**) Number of positive RT-PCR assays, blue columns represent the raw counts of positive assays identified by *S* gene target and red columns the raw counts of *S* dropout (characterized as undetected RT-PCR by *S*, and detected by *ORF1ab* and *N* gene targets). (**B**) Percentage of *S* dropout over epidemiological week with an increase of undetermined results of the *S* gene target on the 10^th^ August week, (8/26, 30.8%, P = 0.007).

All samples with *S* gene dropout eligible for sequencing were from adults, and 24/36 were submitted to WSG. Eight high-quality SARS-CoV-2 whole-genome sequences were recovered (GISAID accession numbers: EPI_ISL_1799498, EPI_ISL_1799499, EPI_ISL_1799501, EPI_ISL_1799502, EPI_ISL_1823203, EPI_ISL_1799504, EPI_ISL_1799505, EPI_ISL_1799507). The other 16 samples were not analyzed, due to the low quality reads and/or low genome coverage. According to the Pangolin COVID-19 Lineage Assigner tool [25] three different lineages were detected: B.1.1.28, B.1.91 and B.1.1.33. Phylogenetic tree corroborated previous results, as shown in Figure 2. Spike protein mutations and INDELs were found, including one mutation (Y380Q) that have never been reported (Table 1).

**Figure 2.**
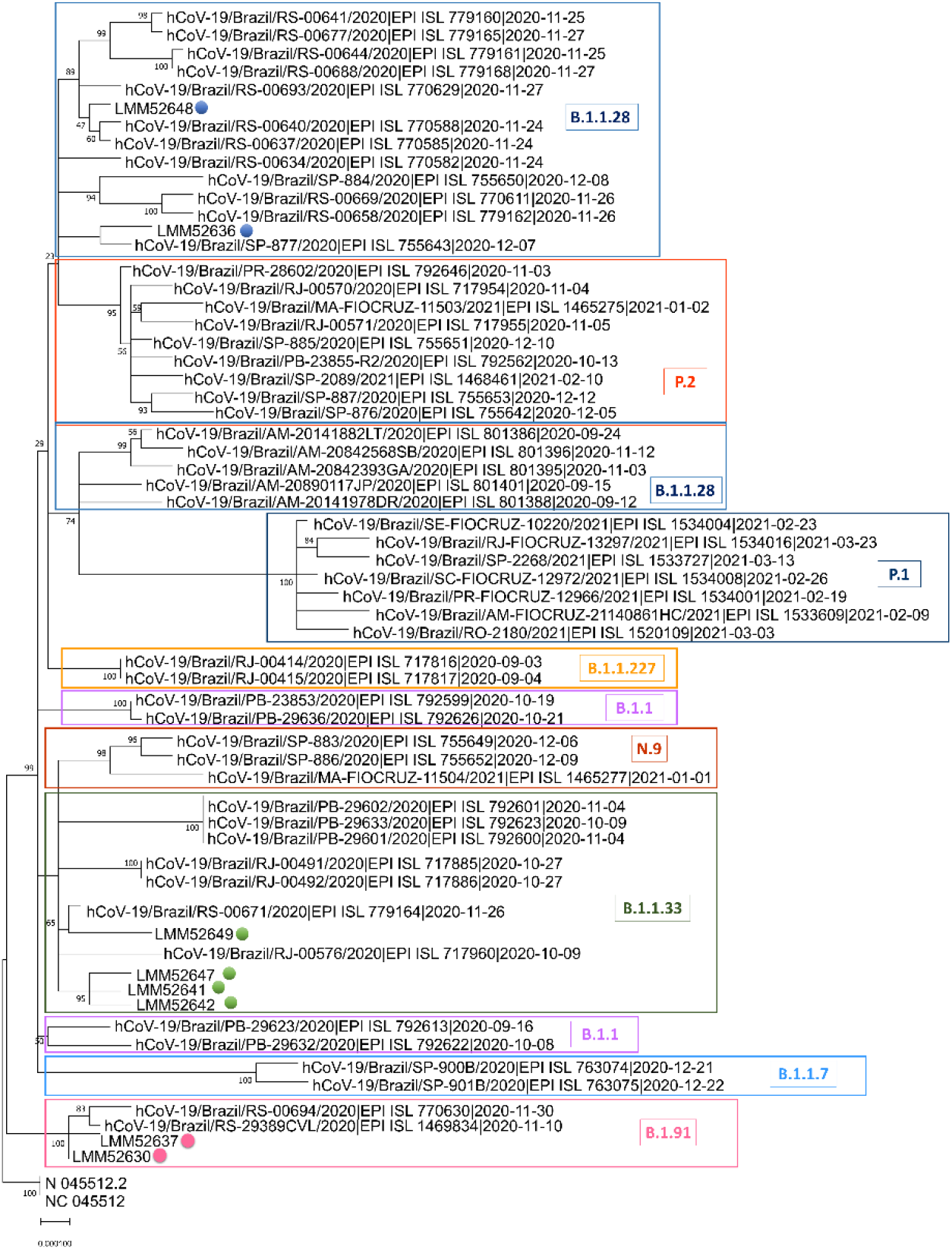
SARS-CoV-2 complete genome phylogenetic tree. The Maximum Likelihood phylogenetic analysis under General Time Reversible allows a proportion of invariable sites, and the substitution rates were inferred empirically in MEGAX web server applying 200 replicates and 1000 bootstrap.

**Table 1.** The consensus and additional mutations, and INDELs observed in the amino acid sequences from eight SARS-CoV-2 infected subjects. In red the novel RBD spike protein mutation.

| ID | Lineage | Consensus mutations of lineages | Additional mutations | INDELs | Date of sample collection |
| --- | --- | --- | --- | --- | --- |
| LMM52630 | B.1.91 | D614G, D839Y | C379W, **Y380Q**, V395A | del:ORF7b:27795:3 | June 2020 |
| LMM52636 | B.1.1.28 | D614G, V1176F | - | - | July 2020 |
| LMM52637 | B.1.91 | G232A, D614G, D839Y, Y1272F | - | - | July 2020 |
| LMM52641 | B.1.1.33 | D614G | T76I, K77N, R78M, F133S | - | August 2020 |
| LMM52642 | B.1.1.33 | D614G | - | del:ORF1ab:686:9 | August 2020 |
| LMM52647 | B.1.1.33 | D614G | F133S | - | August 2020 |
| LMM52648 | B.1.1.28 | D614G, T768N, V1176F | - | - | August 2020 |
| LMM52649 | B.1.1.33 | D614G | - | - | August 2020 |

Median age of the eight sequenced participants was 64.0 years (IQR 39.6-69.3), 75.0% (6/8) were male, and the median days of symptoms onset to inclusion was 7.5 (IQR 2.0-10.0). The most commonly reported symptoms were chills and appetite loss (75.0%), cough, malaise and myalgia (62.5%), and fever (50.0%). Six out of eight participants (75.0%) were hospitalized, 5 (62.5%) required only supplemental oxygen, 3 (37.5%) were admitted to ICU, 2 (25.0%) required mechanical ventilation, and one participant died (1/8, 12.5%). The participant that presented the novel Y380Q mutation was hospitalized for 18 days, including 8-days in ICU, requiring the use of mechanical ventilation.

Wild C379 and Y380 residues of the RBD region were predicted as part B-cell epitopes. NetMHCpan4.1 returns seven binding sequences involving these sites, and two of them were also described as epitopes in the IEDB positive T-cell assays: SASFSTFKCY (for HLA-A^*^01:01 allele) and KCYGVSPTK (for HLA-A^*^03:01 allele). The wild sequence (SASFSTFK**CY**), predicted as a weak binder, turns to a non-binder when mutated (SASFSTFK**WQ**). The wild strong binder sequence (K**CY**GVSPTK) turns into a weak binder (K**WQ**GVSPTK).

The location of identified mutations in the spike protein is depicted in Figure 3A. Structural analysis revealed EP modifications (orange rectangle, Figure 3B) on the LMM52630 models (which includes the B.1.91 lineage model only, and the B.1.91 lineage associated with the RBD substitutions: C379W, V395A and non-reported Y380Q, found exclusively in this participant) compared to the ancestral monomer sequence model. Greater modifications in electrostatic distance (ED = 0.18) are due to the mutations in the B.1.91 lineage (G614 and Y839). Additionally, mutations in RBD presented similar ED = 0.18, shown in the epogram analysis (color bars in Figure 3B). These results can be even more evident examining the surface distribution charges: the D614 and D839 wild residues are negatively charged (red pattern, Figure 3B) and this pattern is disrupted in G614 and Y839 mutated residues; while the surface EP distribution and models conformation show more discreet modifications for the RBD region variants.

**Figure 3.**
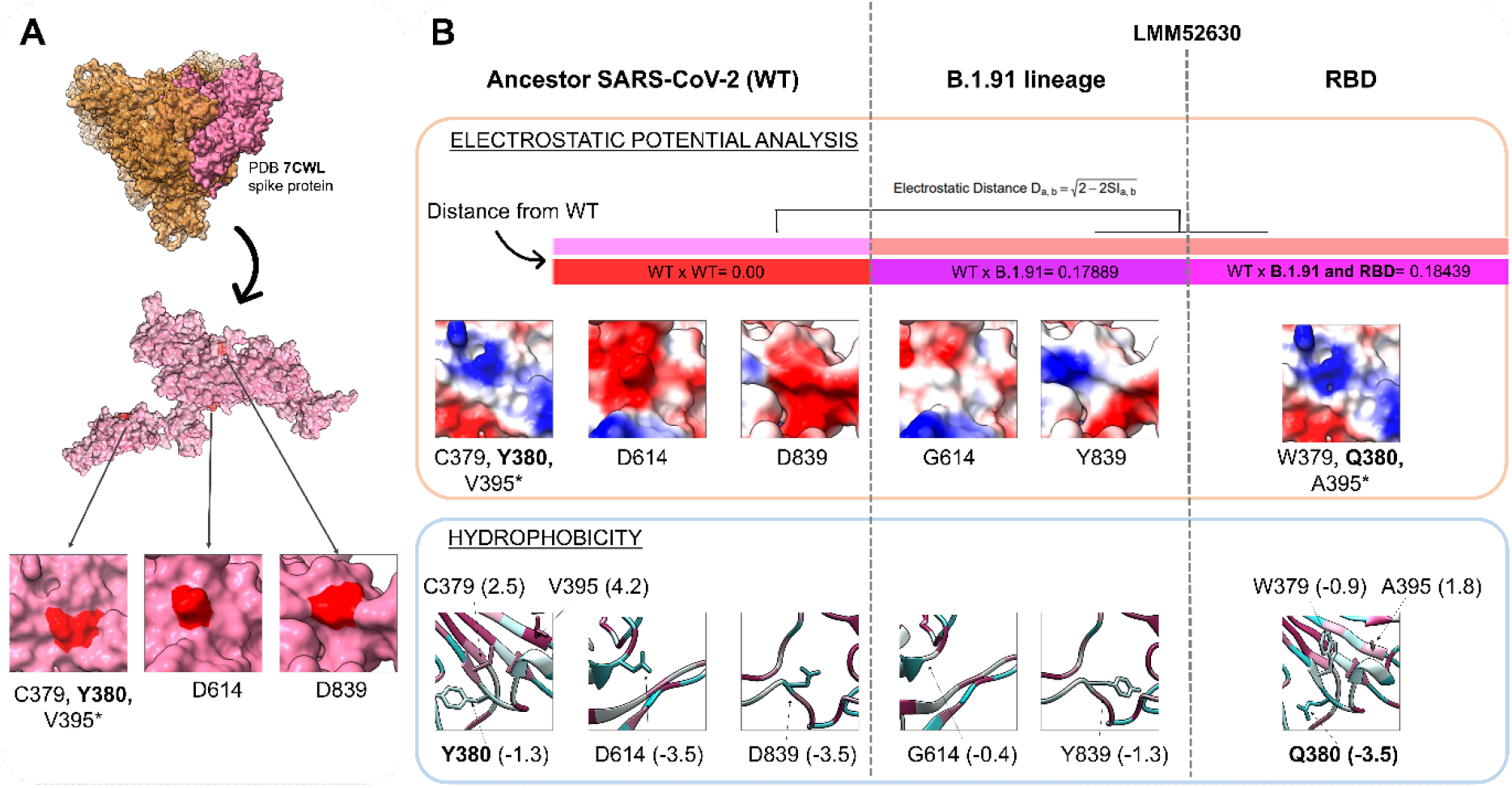
Physicochemical modifications of mutations in the B.1.91 lineage and RBD region (C379W, Y380Q, V395A). (**A**) The PDB 7CWL crystal with the model generated for the spike protein monomer is detached in pink. Variants’ positions at the tridimensional structure surface are highlighted in red. The neighbor residues C379 and Y380 are depicted together, while the V395^*^ is buried and not perceptible at the surface. Both regions (D614 and D839) of the B.1.91 lineage are in exposure regions of the spike protein surface. (**B**) The SARS-COV-2 ancestor lineage (represented by C379, Y380, V395, D614 and D839 residues) was compared to the G614 and Y839 residues of the B.1.91 variant of concern (VOC), and to the mutations presented by the LMM52630 subject (namely: G614 and Y839 of B.1.91 lineage, in addition to W379, Q380 and A395 from RBD residues) considering the electrostatic potential (EP) (orange rectangle) and hydrophobicity (blue rectangle) properties. The epogram (color bars) presents the electrostatic distances (ED) of the mutated models compared to the wild type (ancestor). Both models (B.1.91 lineage and RBD mutations) are similar with ED = 0.18, and diverging from the ancestor. Below of the epogram, are shown the divergences on electrostatic surface distribution, considering the wild and mutated residues. The color scale is represented by a variation from red (more electronegative residues, −5) to blue (more electropositive, +5) passing through neutral (white, 0). The hydrophobicity scale varies from more hydrophobic residues (represented in magenta, positive values) to more hydrophilic (in blue, negative values). (PDB) Protein Data Bank. (RBD) Receptor binding domain. (WT) Wild type.

Amino acid substitutions revealed alterations in hydrophobicity, either by changing the direction of this property (from hydrophobic to hydrophilic and vice versa) or even its intensity. In a general way, substitutions observed in the B.1.91 lineage turn their regions to more hydrophobic, while the RBD mutations to more hydrophilic, denoted by negative and positive values (blue rectangle, Figure 3B) considering hydrophilic and hydrophobic patterns, respectively. Substitutions from wild residues D614 (−3.5) and D839 (−3.5) to G614 (−0.4) and Y839 (−1.3) incremented the hydrophobicity in B.1.91 lineage, while the substitution of Y380 (−1.3) and V395 (4.2) to Q380 (−3.5) and A395 (1.8) reduced this property. The C379W mutation changed the direction from highly hydrophobic (2.5) to hydrophilic (−0.9). Altered residues in the RBD region, specially the C379W and Y380Q mutations, are located close to each other and likely gain strength, thus providing an overall shift to hydrophilic profile. The changing potential of these two close mutations induced to the buried A395 a more hydrophilic profile when compared to the ancestor (from 4.2 to 1.8).

A structural investigation of more than 20 crystals of viral spike protein from PDB, revealed that the mutations C379W, Y380Q and V395A are in a contact area complexed with antibodies in the RBD region. And four crystals that presented Fab antibodies (fragment antigen-binding) are in contact with 379 and 380 RBD residues. Figure 4B exhibits the CR3022 human antibody complexed with the RBD region in contact with mutation sites. We observed the same when evaluating the S2H97, EY6A and S304 antibodies. H-bonds are important non-covalent interaction forces which can assist in protein residue bindings, especially in stabilizing antibody-antigen interactions [26]. Mutated Q380 residue leads to an H-bond disruption observed previously between the wild Y380 with the S99 residue of the CR3022 neutralizing antibody. While the W379 alteration disables the previous H-bond between the wild C379 with T94 residue of the EY6A neutralizing antibody. Mutated W379 also affects the H-bond neighboring of the G381 with Y92 residue of EY6A antibody. RBD-antibody interaction sites are shown in Supplementary Table 1.

**Figure 4.**
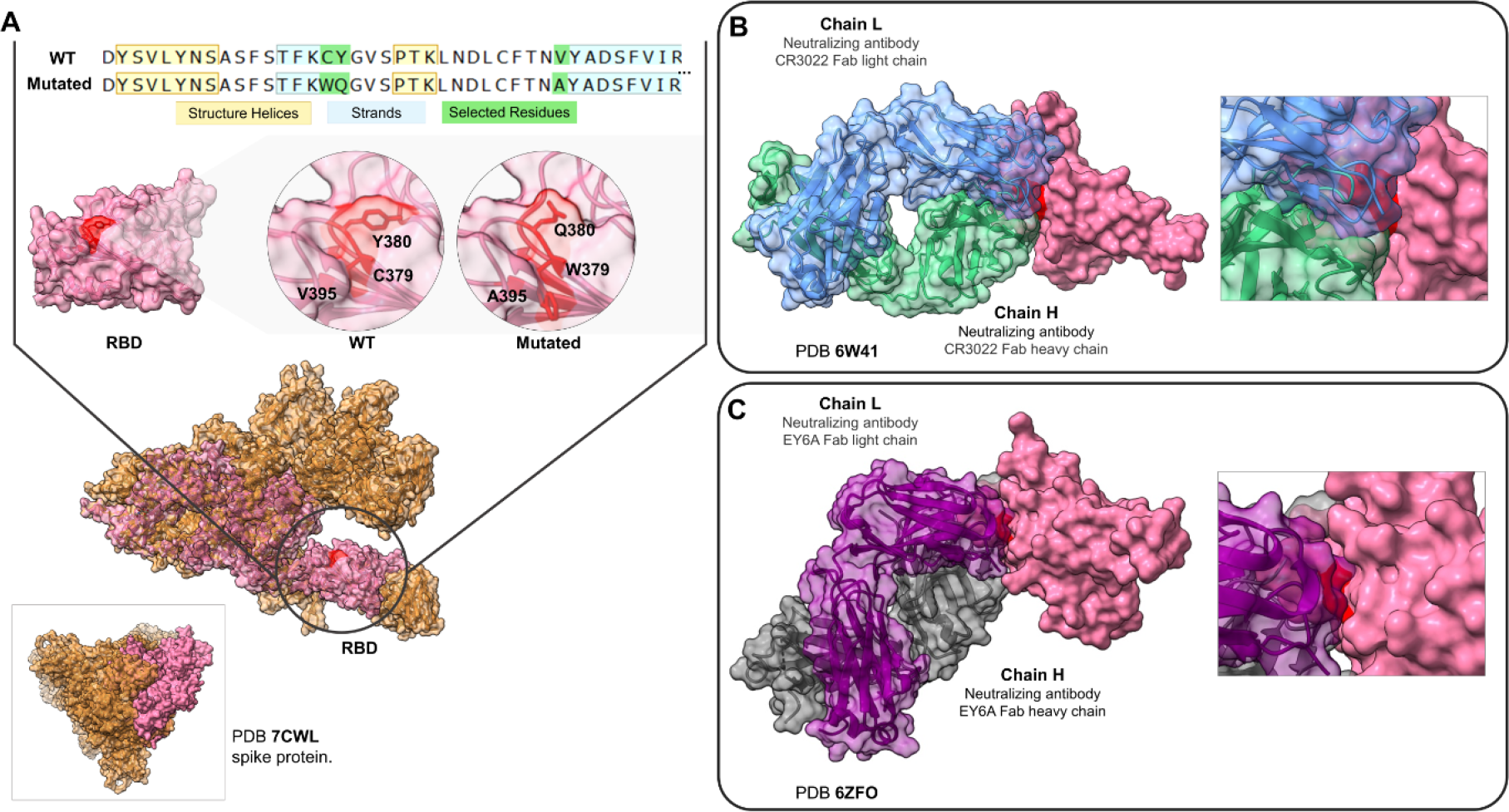
RBD mutation regions of the LMM52630 subject in contact with neutralizing antibodies. (**A**) The comparison of the mutated sequences with the wild type. Below, the structural model with the selected residues, depicted (red color) for wild and mutated RBD region. The model of the monomer was obtained from the 7CWL PDB structure. (**B**) The contact region among the RBD (pink), the Fab CR3022 neutralizing antibody (light and heavy chains, represented in blue and green colors, respectively) and the residues C379 and Y380 (red). (**C**) The same RBD region contacting the neutralizing antibody EY6A (the light chain in purple and heavy in grey). (RBD) Receptor binding domain. (WT) Wild type.

## Discussion

Our results suggest that SARS-CoV-2 *S* gene dropouts were present in the community as early as in the beginning of the pandemic in Southern Brazil. These results may indicate that the spread of mutations, resulting in different genetic variants of the virus, were already circulating much earlier than recognized in most settings. Further, a new mutation (Y380Q) was identified, and together with C379W modify important properties in the RBD region, which may interfere with the binding of neutralizing antibodies (CR3022, EY6A, H014, S304).

*S* gene dropout has been found strongly associated with a six-nucleotide deletion resulting in the loss of two amino acids: H69 and V70. Despite the WGS, we could not directly link the *S* dropout to specific viral mutations. This result may be due to the small sample size submitted to sequencing. Though, we found the presence of three different lineages B.1.1.28, B.1.91 and B.1.1.33. Additionally, the Y380Q mutation was identified in the *S* gene, and thus could be associated with the natural history of virus evolution or may possibly be associated with the emergence of new and more virulent mutations.

It is well known that modifications in the EP and hydrophobicity distribution may interfere in protein-protein interaction. Interface regions are usually composed of residues presenting opposite charges and hydrophobic pairs, where small changes on these properties, in important functional sites, may impact canonical interactions [27]. Neutralizing antibodies (NAbs) are fundamental elements of the immune system against viral infections [28], and the H-bonds of wild C379 and Y380 residues with the S304, CR3022, S2H97 and EY6A NAbs reinforce the importance of these regions. Therefore, the H-bond disruptions observed in W379 and Q380 substitutions plus the alteration in hydrophobicity disfavor the RBD-antibodies interactions. As the C379 residue is part of one of the four disulfide bonds in the RBD region (C379-C432) its disruption could generate instability since it contributes to β sheet conformation maintenance [29].

A previous study reported that the C379 and Y380 residues are part of an epitope for H014 antibody, which could sterically compete with the ACE2 host molecule for the RBD interaction [30]. It also reported an overlap among the binding epitopes for the H014 and CR3022 antibodies. The CR3022 monoclonal antibody neutralizes the RBD region of SARS-CoV-2, disrupts the prefusion spike conformation, and also competes sterically with ACE2 [31] without physically blocking it [32]. The recently described S2H97 is a potential NAb [33] that exhibits a notable tight binding even with divergent RBD regions from other Sarbecoviruses. Other SARS-CoV-2 NAbs that bind to the RBD were also described as non-overlapping with the ACE2 binding site [34].

This study has some limitations. The small sample size in which WGS was feasible may limit any conclusion about the clinical severity related to the mutations found. Moreover, individuals were enrolled in a single city in Southern Brazil, which may limit the generality of our findings. Nonetheless, despite such limitations, a novel mutation (Y380Q) in the RBD region of SARS-CoV-2 spike protein was described. The analysis based on crystal structures reinforces the importance of the Y380 and C379 residues in the NAbs binding, and thus mutations in these regions may affect the interaction effectiveness between the NAbs and SARS-CoV-2 protein, as inferred by computational analysis.

## Conclusions

Our findings indicate that SARS-CoV-2 variants were circulating quite early in the community. A possible role of the new described mutations with clinical severity can be speculated, but further studies are needed to confirm this hypothesis. Studies assessing the mutations and their relation to prognosis are necessary, and also to evaluate vaccine effectiveness in a challenging scenery that is continuously changing.

## Data Availability

All data are available in the manuscript and in the supplementary material.

## Supplementary material

**Supplementary Figure 1.**
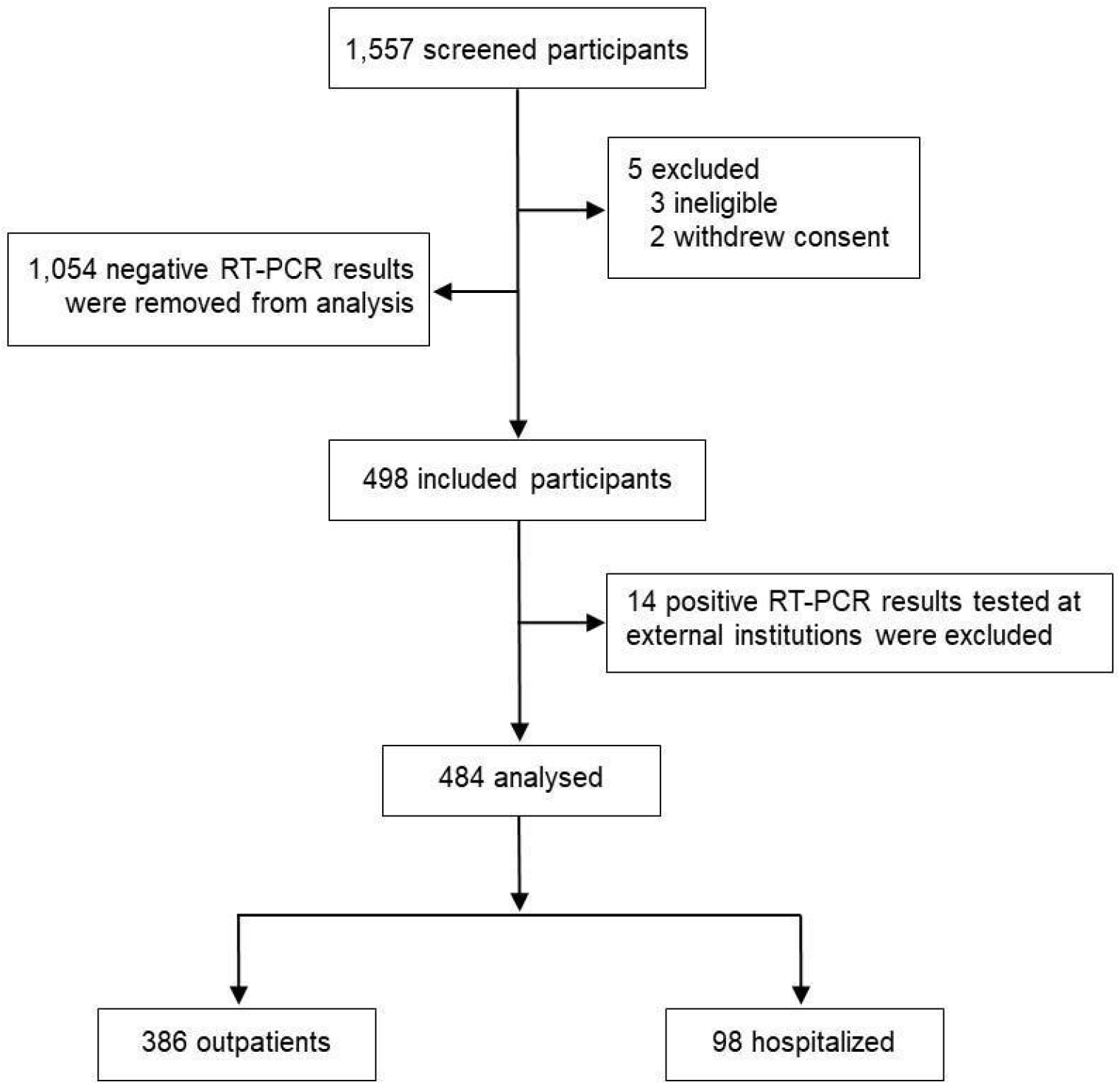
Subjects flowchart. 1,557 subjects were screened and 19 were excluded (3 for not meeting inclusion criteria and 2 withdrew consent; 14 were positive for SARS-CoV-2 infection but were tested at external institutions), and 1,054 participants with negative RT-PCR results were removed from analysis. A total of 484 participants with SARS-CoV-2 infection confirmed by RT-PCR were analyzed. Of this total, 386 were outpatients and 98 inpatients.

**Supplementary Table 1.** The wild RBD-antibody interaction sites and the mutated models.

| Antibody | PDB code | RBD residues with direct contact to antibody |
| --- | --- | --- |
| CR3022 | 6W41 | Y369, N370, S371, **F377(Y52-H)**, **K378(D54-H, E56-H)**, **C379(I98-H)**, **Y380(S99-H)**, **G381(Y32-L)**, V382, S383, P384, T385, **K386(D101-H)**, L390, F429, **T430(S27-L)**, F515, E516, L517 |
| CR3022 | Mutatted model | Y369, N370, S371, **F377(Y52-H)**, **K378(D54-H, E56-H)**, **W379(I98-H)**, **G381(Y32-L)**, V382, S383, P384, T385, **K386(D101-H)**, L390, F429, **T430(S27-L)**, F515, E516, L517 |
| EY6A | 6ZFO | Y369, F377, **C379(T94-L)**, Y380, **G381(Y92-L)**, **V382(T94-L)**, S383, P384, **T385(N33-H, Y106-H)**, **N388(Y53-H)**, L390, F392, P412, D427, **F429(Y92-L)**, L517 |
| EY6A | Mutated model | Y369, F377, **C379(T94-L)**, Y380, **V382(T94-L)**, S383, P384, **T385(N33-H, Y106-H)**, **N388(Y53-H)**, L390, F392, P412, D427, **F429(Y92-L)**, L517 |
| S304 | 7JW0 | Y369, F377, K378, **C379(S94-L1)**, Y380 |
| S2H97 | 7M7W | Y369, N370, S371, A372, F374, S375, K378, **C379(Y105-H)**, Y380, V382, S383, P384, T385 |

The RBD region performing hydrogen bonds with antibodies are shown in bold. The residue chains of the antibody sequence are depicted as light (L) or heavy (H) after the antibody residue. In red are denoted the wild type and mutated 379 and 380 residues.

## Acknowledgments

We thank the Scientific Committee of the Research Support Nucleus (Núcleo de Apoio à Pesquisa, NAP) of Hospital Moinhos de Vento for technical-scientific consultancy. We thank the inclusion personal, laboratory team, and site staff from Hospital Moinhos de Vento and from Hospital Restinga e Extremo Sul. Aline Andrea da Cunha, Joao Ronaldo Mafalda Krauser, Paulo Sergio Kroeff Schmitz, Sidiclei Machado Carvalho, Fabio Jose Rockenbach, Thaís Pacheco Oliveira, Cláudia Josiel Oliveira do Coito, Kelly Viegas Antunes, Marcelo da Silva Ferreira, Rafael Garcia Trindade, Thayna Silva Lino, Tiago da Silva Silvano, Adriana da Silva Silveira, Alceu Kuckoski, Ana Paula da Silva Lopes, Andreia Escobar da Costa, Clarice Cardoso Machado, Erica Vieira da Silva, Evelin Inácia da Silva, Luciana Rodrigues Ribeiro, Marcely Mayr da Costa, Morgana Thais Carollo Fernandes, Rafael da Silva Cassafuz.

## COVIDa study group

Adriane Isabel Rohden, Ana Paula dos Santos, Camila Dietrich, Caroline Cabral Robinson, Catia Moreira Guterres, Débora Vacaro Fogazzi, Denise Arakaki-Sanchez, Fernanda Lutz Tolves, Fernando Rovedder Boita, Francieli Fontana Sutile Tardetti Fantinato, Gisele Alcina Nader Bastos, Jaina da Costa Pereira, Maicon Falavigna, Maristênia Machado Araújo, Patricia Bartholomay Oliveira, Regis Goulart Rosa, Shirlei Villanova Ribeiro, Thainá Dias Luft, Tiago Fazolo.

